# First report of genetic variants detected in Argentinian patients with clinical Long QT Syndrome diagnosis

**DOI:** 10.1101/2023.02.15.23285915

**Authors:** Dionisio Leonardo, Stupniki Sofía, Aztiria Eugenio, Rías Ezequiel, Dye Leandro, Onetto Leonardo, Gregorietti Franco, Keegan Roberto, Spitzmaul Guillermo

## Abstract

**Background:** Long QT Syndrome (LQTS) is a genetic cardiac condition in which disease severity and response to pharmacological treatments vary according to genetic variations. In Argentina, most of the LQTS diagnoses are made by clinical exploration and ECG analysis. In this work, we evaluated a group of subjects from our community to correlate their clinical LQTS diagnosis with genetic modifications. *Material and methods*: Using gDNA isolation, PCR, and exome sequencing, we screened the coding sequences of the *KCNQ1, KCNH2*, and *SCN5A* genes in the studied cohort.

**Results:** We identified several DNA changes, among synonymous and non-synonymous, most of them previously described in the literature. In addition, we found a non reported alteration in the sequence of *KCNQ1* sequence that suggests the lack (deletion) of an exon or a large part of it indicating exon deletion. 16 which did not allow us to amplify it.

**Conclusions:** This is the first report of genetic variations in LQTS-associated genes in Argentina. The variations detected could explain the prolongation of the QT interval observed in the ECG of some of the individuals or those with a suspicious family history and could improve treatment, making it more rational as well as providing genetic counselling to first-degree relatives.

**Highlights:**

- Genetic screening correlates with clinical diagnosis in LQT patients
- Studied cases carry more than one variation in at least 2 genes simultaneously
- A Non-reported variation in KCNQ1 exon 16 was founded in one case

## 1. Introduction

Long QT syndrome (LQTS) is a congenital genetic disorder that can cause lethal cardiac arrhythmia and sudden cardiac death^1^. Patients with LQTS display a prolonged corrected QT interval (QTc) on resting ECG (i.e. >440 ms in men and >460 ms in women). The prevalence of this disease in the world is about 1 in 2,000 persons^2^. Since it is a rare condition, there are no official statistics in Argentina. About 15 genes encoding ionic channels or associated proteins have been implicated in this disease. The most frequently involved are *KCNQ1, KCNH2*, and *SCN5A*, which encode for the slow rectifying K^+^ channel KCNQ1, the fast rectifying K^+^ channel hERG, and the fast Na^+^ channel Nav1.5, respectively^3^. Dysfunction of one or more of these channels may lead to lengthening the QT interval in ECG. About 80% of the patients with clinically diagnosed LQTS have mutations in one of these 3 genes: *KCNQ1* (40-45%), *KCNH2* (30-35%), and *SCN5A* (10%)^2^ which classify the disease in 3 major subtypes LQT1, LQT2, and LQT3, respectively. In addition, between 5-10% of the patients with LQTS carry several alterations in more than 1 of these genes simultaneously. These patients generally have a more severe phenotype and earlier onset^4^.

QT interval prolongation is a pathognomonic sign of the disease, but up to one-third of mutation carriers may have normal QT intervals on resting ECGs^3^. This is why genetic testing for mutations that lead to cardiac channelopathies is essential for an accurate diagnostic evaluation and familial screening. Molecular identification of the causes of this disease contributes not only to a better diagnosis but also to risk stratification, genetic counseling, and treatment of this population.

Our aim in this study was to correlate the clinical diagnosis with genetic variants of LQTS. Furthermore, to understand the spectrum of genetic variations that underlie this condition in Argentinian subjects, we examined LQTS cases, including patients and relatives, for alterations in the three most frequently implicated LQTS genes, *KCNQ1, KCNH2*, and *SCN5A*, by exon amplification and direct DNA sequencing.

## 2. Materials and methods

### 2.1. Ethics statement

Written informed consent for genetic explorations and authorization for publication of the results of this study were obtained from patients and family members. This study was conducted under the approval of the ethics committee of the *Hospital Municipal de Agudos L. Lucero* de Bahía Blanca (protocol #UNS8614) following the Declaration of Helsinki.

### 2.2. Case selection

The Cardiac Electrophysiology Service of the *Hospital Privado del Sur of Bahia Blanca (Argentina)* selected the cases with a clinical diagnosis of LQTS to perform the molecular analysis. This selection was carried out following the Schwartz criteria^5^. We investigated 6 Argentinian individuals (3 men and 3 women) with an age range that went from the first year of life to 62 years. As a control population, we studied 5 healthy unrelated individuals of the same origin, with an age range between 20 and 60. In the first group, 5 out of the 6 individuals, showed a prolonged QTc interval on the ECG (i.e. >460 ms) while one of them (case #2 in Table 1) was a first-degree relative presenting a normal QTc interval (<450 ms). For case #5, we added 2 first-degree relatives (sister and father) to evaluate a specific variation.

**Table 1.** Clinical features of individuals with clinical LQTS diagnosis.

| Case | Sex | Age range | Presentation | Family history | Arrhythmia | QTc (ms) | Therapy |
| --- | --- | --- | --- | --- | --- | --- | --- |
| 1 | F | 0-5 | asymptomatic | None | None | <450 | None |
| 2 | M | 26-30 | asymptomatic | None | None | 510 | BB |
| 3 | F | 30-35 | symptomatic | None | Yes | 460 | BB + CA + ICD |
| 4 | M | 40-45 | symptomatic | None | Yes | 495 | BB |
| 5 | M | 6-10 | asymptomatic | Yes | None | 500 | BB |
| 6 | F | 60-65 | symptomatic | None | None | 480 | BB |

M: male, F: female, BB: beta-adrenergic receptor blocker, CA: catheter ablation, ICD: Implantable cardioverter-defibrillator.

### 3.3. PCR amplification

Genomic DNA (gDNA) was extracted from whole blood cells using a commercial kit (Inbio Highway, Argentina). gDNA was used for PCR amplification of coding exons of *KCNQ1, KCNH2*, and *SCN5A*. The *KCNQ1* gene has 16 coding exons, the *KCNH2* gene has 15 and the *SCN5A* gene has 28. Specific primers were designed to amplify each exon (Tables 2, 3, and 4). Each primer pair was designed to hybridize ∼100 bp before and after the coding region. For the KCNH2 gene, exon 1 codifies for 25 aminoacids however few polymorphisms were reported in it, so it was not amplified. For the SCN5A gene, exon 1 is not a codifying region, so it was also not amplified. Optimal PCR conditions were set up empirically for each reaction. PCR products were separated by agarose gel electrophoresis and purified using a commercial kit (PB-L, Argentina).

**Table 2.**
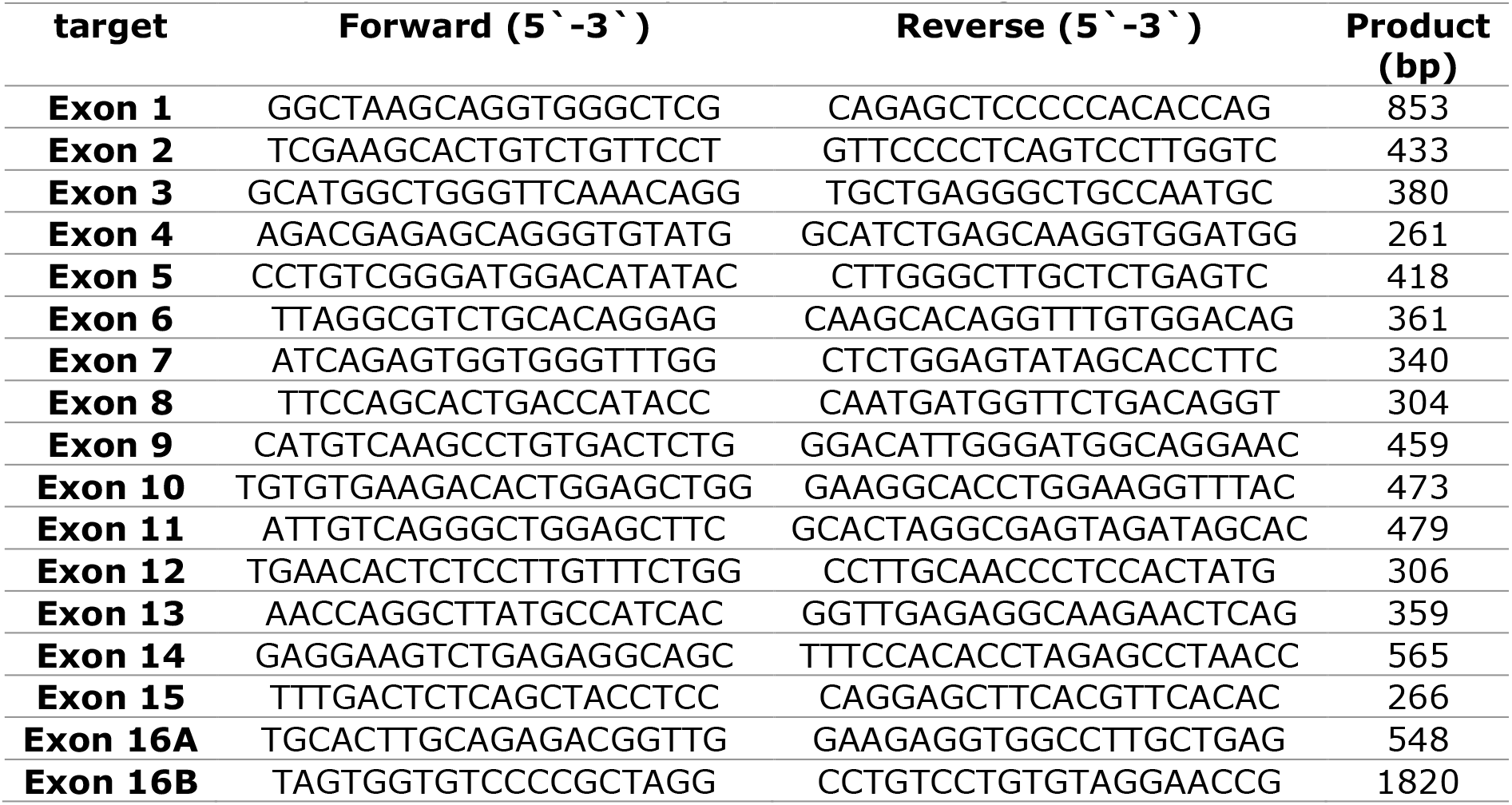
List of primers used to amplify KCNQ1 coding exons.

**Table 3.**
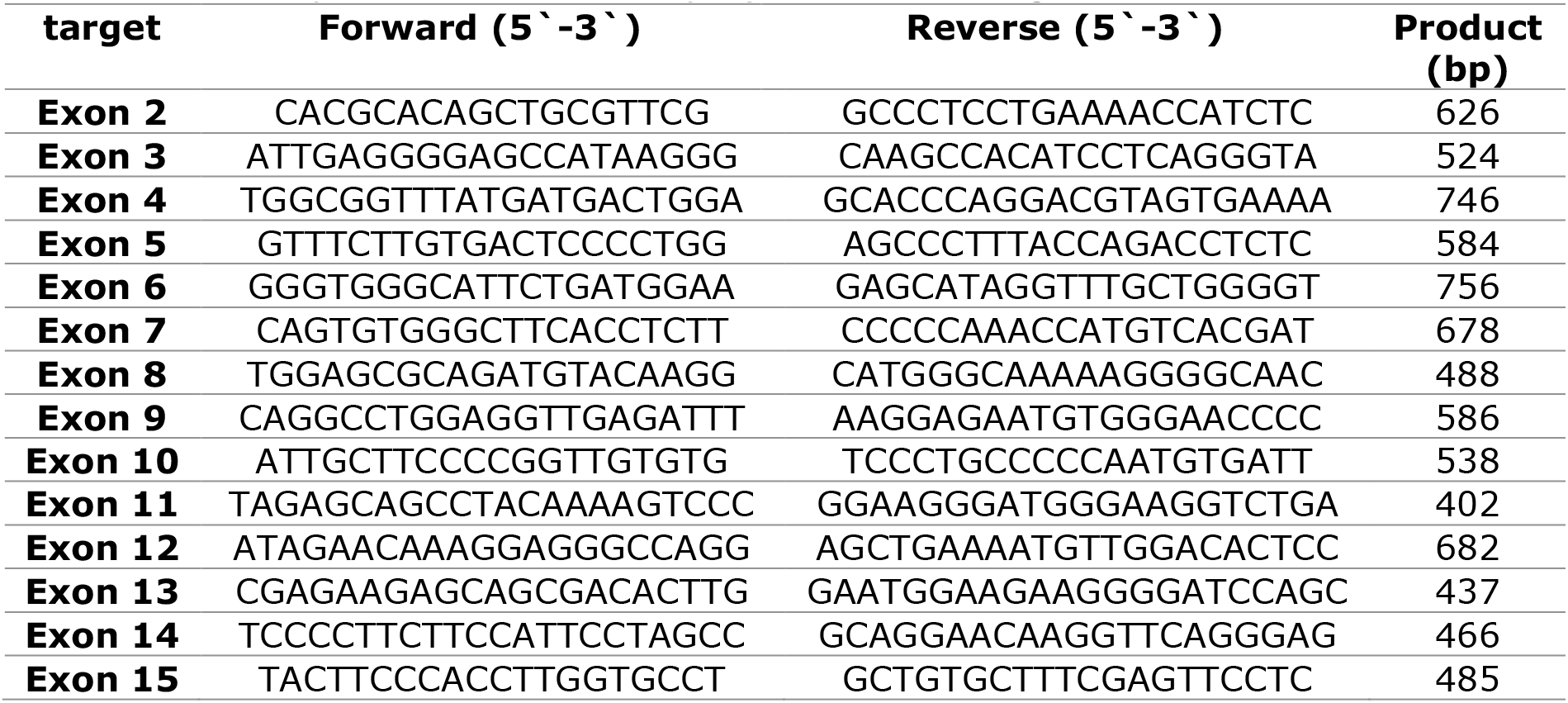
List of primers used to amplify KCNH2 coding exons.

**Table 4.**
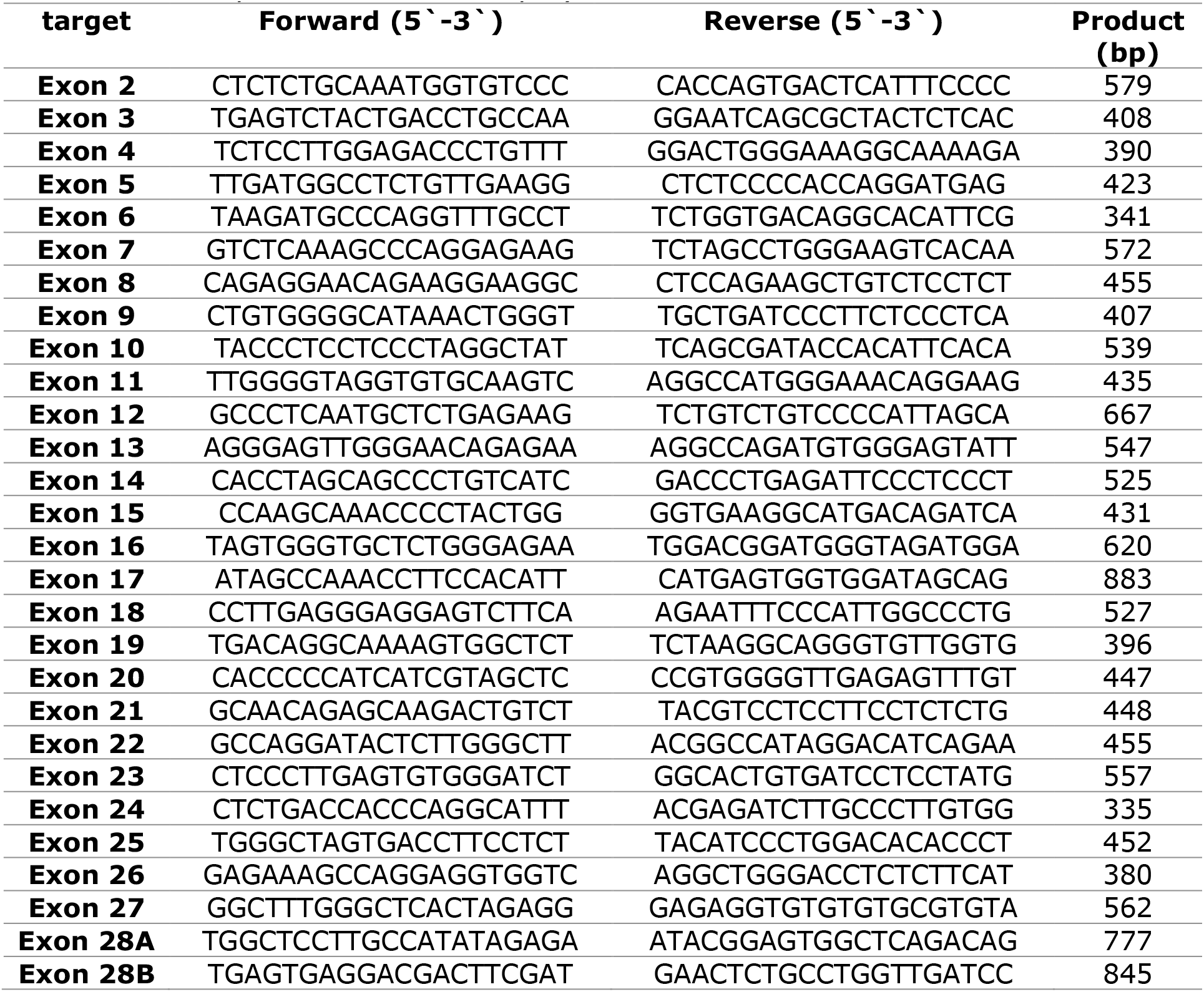
List of primers used to amplify SCN5A exons.

### 3.4. DNA sequencing and analysis

PCR products were sequenced in both directions (5’→3’and 3’→5’) using an external service (Macrogen Inc., Korea). DNA alignments and polymorphism detection were carried out offline using software programs such as SnapGene (V5.2.4) and Benchling. The sequences of the complete human *KCNQ1* (Accession Number NG_008935.1), *KCNH2* (Accession Number NG_008916.1), and *SCN5A* (Accession Number NG_008934.1) genes described in GenBank were used as reference.

### 3.4 Structural analysis of human KCNQ1, hERG, and Nav1.5

Homology modeling was performed using the SWISS-MODEL server (https://swissmodel.expasy.org)^6^. The 3D protein model was generated employing as target protein the FASTA amino acid sequences for human KCNQ1 (accession number NP_000209.2), hERG (NP_000229.1), and Nav1.5 (NP_932173.1).

## 4. Results

### 4.1 Clinical features

In the group of individuals selected for the study, 5 out of 6 of them, showed a prolonged QTc interval on the ECG (i.e. >460 ms) while the remaining one (case #2 in Table 1) was a first-degree relative presenting a normal QTc interval (<450 ms). Typical ECG recordings of each subject are shown in Fig 1. One of the patients had a history of sudden cardiac death before the age of 40, and another had presyncope and documented polymorphic ventricular tachycardia (see Table 1). Some of the subjects are under pharmacological treatment with β-adrenergic receptor blockers (indicated in table 1).

**Fig. 1:**
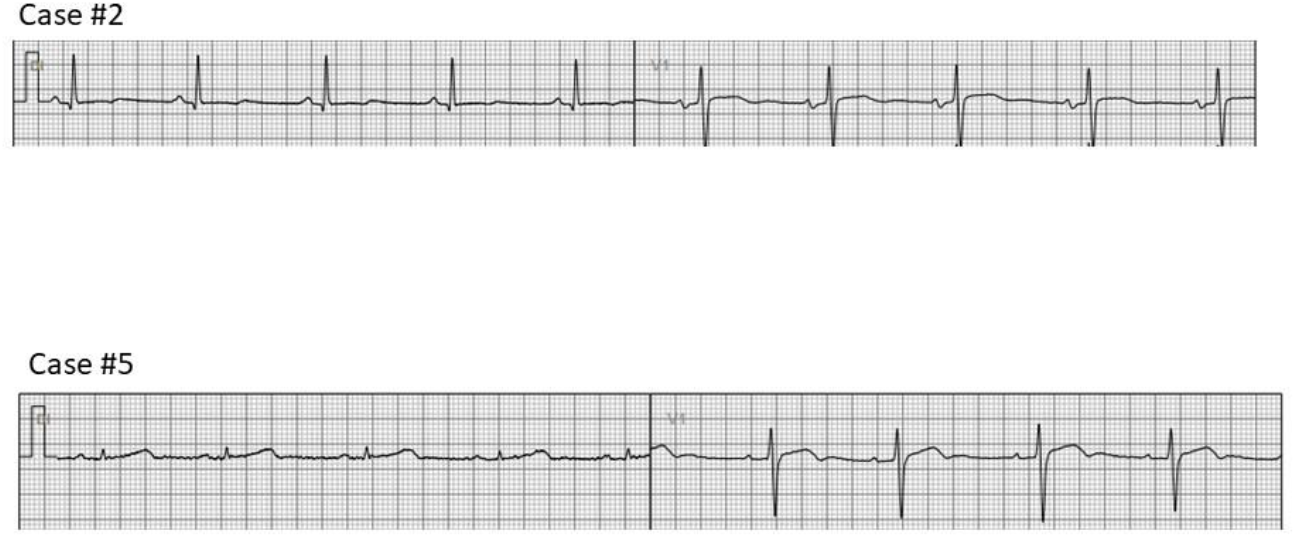
Examples of ECG of two of the studied individuals.

### 4.1. *KCNQ1* sequencing analysis

The results of the molecular screening of the *KCNQ1* gene in the different cases are presented in Table 5 and Fig 2.

**Table 5.** Detected variants for 6 cases of the studied cohort.

| Case | Gene | Region | Nucleotide Change | Amino acid change | Zygosity | ClinVar |
| --- | --- | --- | --- | --- | --- | --- |
| #1 | KCNQ1 | - | - | - | - | - |
|  | KCNH2 | Exon 7 | c.1692A>G | p.L564L | Htz | B |
|  |  | Exon 8 | c.1956T>C | p.Y652Y | Htz | B |
|  | SCN5A | Exon 17 | c.3183A>G | p.E1061E | Hmz | B |
| #2 | KCNQ1 | Exon 13 | c.1638G>A | p.S546S | Htz | B |
|  | KCNH2 | Exon 8 | c.1956T>C | p.Y652Y | Htz | B |
|  |  | Exon 11 | c.2690A>C | p.K897T | Htz | B/LP |
|  | SCN5A | Exon 2 | c.87A>G | p.A29A | Htz | B |
|  |  | Exon 17 | c.3183A>G | p.E1061E | Hmz | B |
|  |  | Exon 28 | c.5454C>T | p.D1818D | Htz | B |
| #3 | KCNQ1 | - | - | - | - | - |
|  | KCNH2 | Exon 5 | c.982C>T | p.R328C | Htz | P |
|  | SCN5A | Exon 12 | c.1673A>G | p.H558R | Htz | B/LP |
|  |  | Exon 17 | c.3183A>G | p.E1061E | Hmz | B |
| #4 | KCNQ1 | - | - | - | - | - |
|  | KCNH2 | Exon 8 | c.1956T>C | p.Y652Y | Hmz | B |
|  | SCN5A | Exon 2 | c.87A>G | p.A29A | Hmz | B |
|  |  |  | c.100C>T | p.R34C | Htz | B |
|  |  | Exon 12 | c.1673A>G | p.H558R | Htz | P/LP |
|  |  | Exon 28 | c.5454T>C | p.D1818D | Htz | B |
|  |  |  | c.5841C>T | p.I1947I | Htz | B |
| #5 | KCNQ1 | Exon 16 | Not amplified | - | - | n. d. |
|  | KCNH2 | Exon 8 | c.1956T>C | p.Y652Y | Hmz | B |
|  | SCN5A | Exon 2 | c.87A>G | p.A29A | Htz | B |
|  |  | Exon 17 | c.3183A>G | p.E1061E | Hmz | B |
| #6 | KCNQ1 | Exon 13 | c.1638G>A | p.S546S | Htz | B |
|  | KCNH2 | Exon 8 | c.1956T>C | p.Y652Y | Hmz | B |
|  |  | Exon 11 | c.2690A>C | p.K897T | Hmz | B/LP |
|  | SCN5A | Exon 2 | c.87A>G | p.A29A | Htz | B |
|  |  | Exon 5 | c.535C>T | p.R179* | Htz | P |
|  |  | Exon 17 | c.3183A>G | p.E1061E | Hmz | B |

*n. d.*: not described, *Hmz*: homozygous, *Htz*: heterozygous, *ClinVar*: ClinVar variant interpretation category, *B*: benign, *P/LP* pathogenic/likely pathogenic, *P*: pathogenic.

**Fig. 2.**
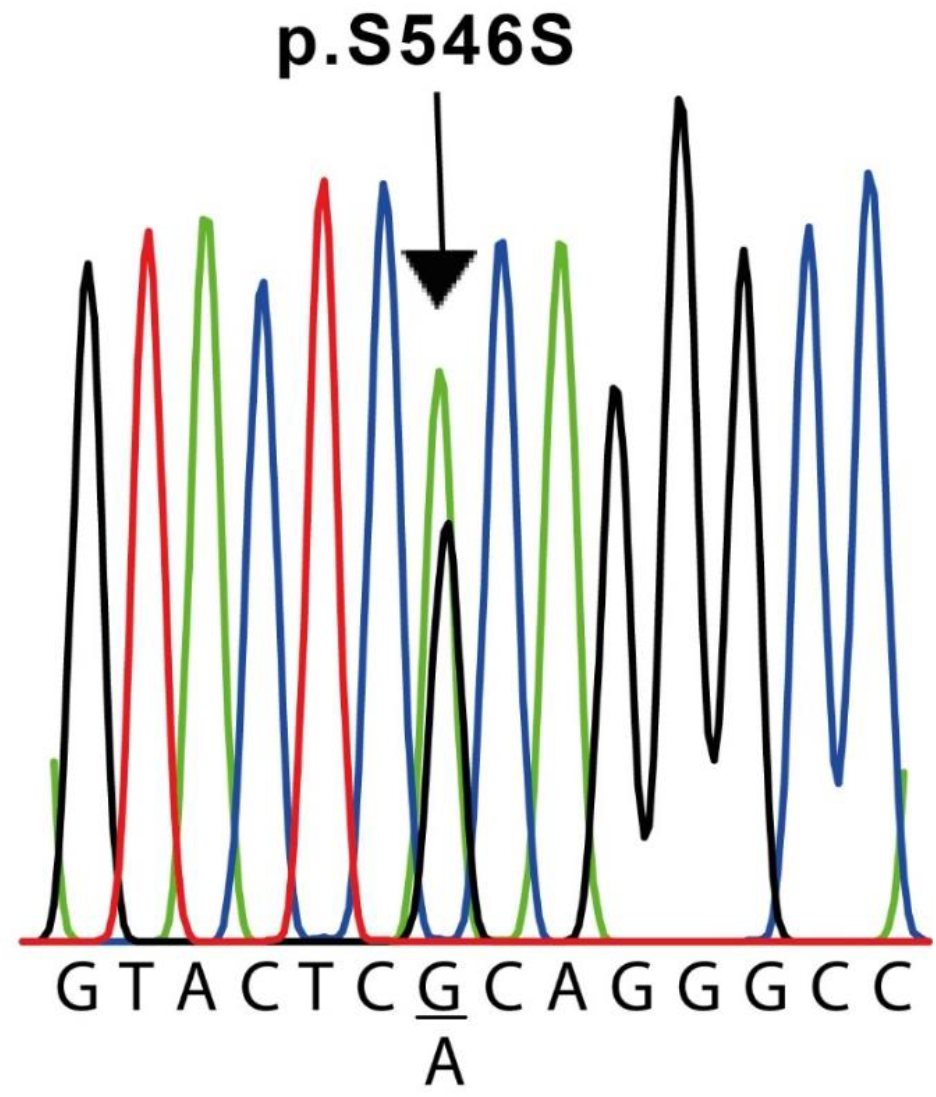
Nucleotide sequence of the KCNQ1 variant found in cases #2 and #6. Both patients underwent a c.1638G>A exchange in exon 13 in a heterozygous configuration, resulting in a silent mutation (p.S564S). For the sake of clarity, the sequence shown is only for the sense strand of patient #2 genomic DNA. The complete sequence of the human KCNQ1 gene described in GenBank (Accession Number NG_008935.1) was used as a reference.

Exons 2 to 16 of this gene were analyzed in all 6 patients and the control subjects. On the other hand, it was not possible to amplify exon 1 in any sample. We identified 1 polymorphism of the *KCNQ1* gene and 1 change in the genomic sequence (Table 5). For this gene, we detected the c.1638G>A (p.S546S) variant in exon 13 of cases #2 and #6 (Fig. 2). In both cases, this variant was found in heterozygosis. Additionally, in our experimental conditions, we were unable to amplify exon 16 of case #5 but on the contrary, we were able to do it in the other five cases and the five control samples. To verify this result, two different samples from the patient, obtained independently, were used. To exclude the possibility that the original pair of primers would not anneal correctly on the target sequence, we designed a second pair of primers located far before and after in the sequence (around 600 bases away from the original pair of primers, see table 1). Also, in this case, we were unable to amplify this exon in this individual. To verify the presence of the same variant in 2 first-degree relatives (father and sister), we performed the same PCR reaction for exon 16 in both. In these two individuals, the reaction allowed to correctly amplify the exon in question.

### 4.2 KCNH2 sequencing analysis

The results of the molecular screening of the *KCNH2* gene are presented in Table 5. All coding exons of this gene were analyzed in the selected population and the corresponding controls. We identified 5 polymorphisms of the *KCNH2* gene (Table 5). Among these there were the c.1692A>G (p.L564L) variant in exon 7 of case #1, the c.2690A>C (p.K897T) variant in exon 11 of case #2 and #6, the c.982C>T (p.R328C) variant in exon 5 of case #3, the c.35951C>A (p.A1077A) variant in exon 14 of case #5. In addition, we detected the c.1956T>C (p.Y652Y) genomic variant in exon 8 of all cases except case #3 (Table 5 and Fig. 3). All of them located at the transmembrane and intracellular domains (Fig. 5).

**Fig. 3.**
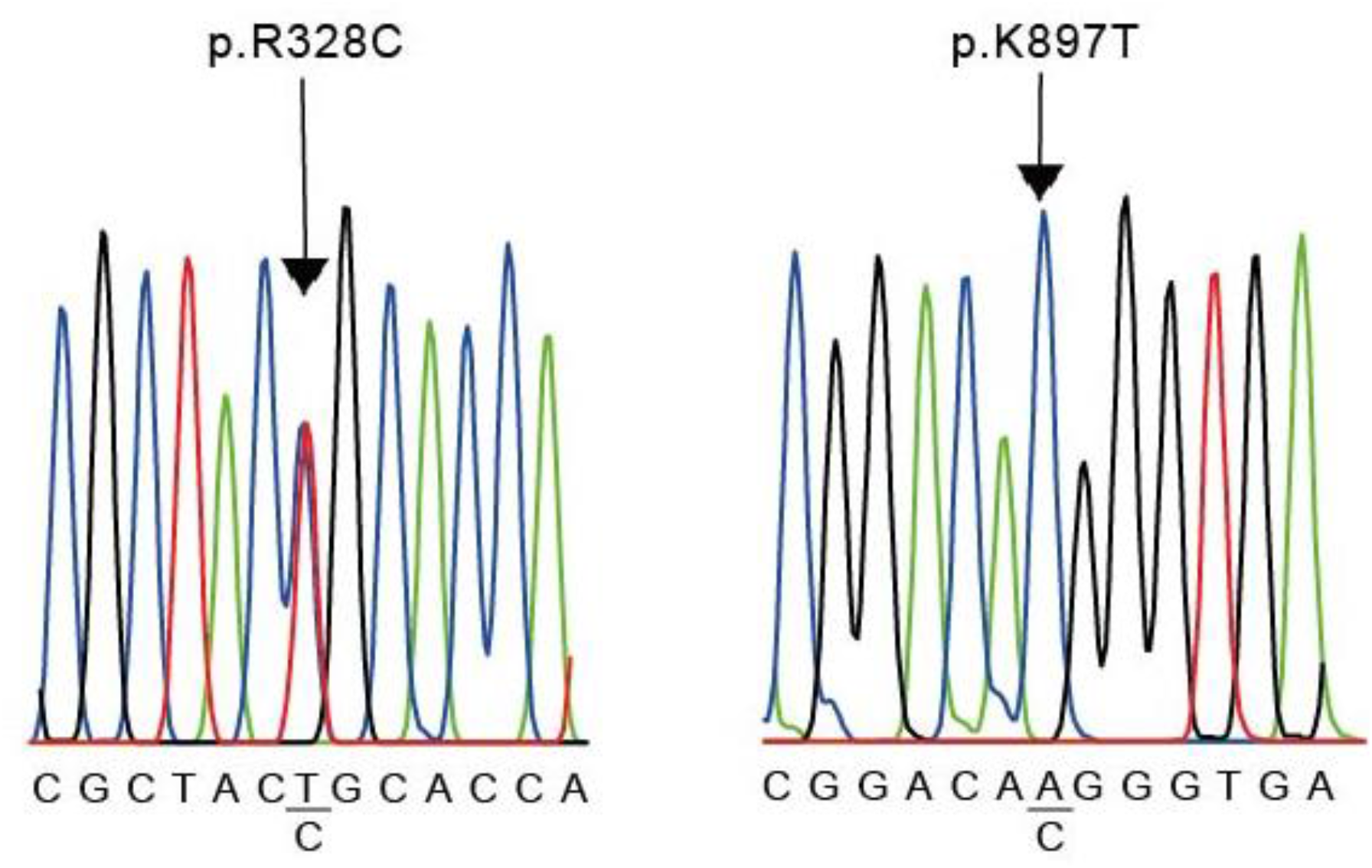
Nucleotide sequence of KCNH2 variants. R238C was founded in case #3 (left) and K897T was found in cases #2 and #6 (right). The sequence shown is that of the sense strand of patients genomic DNA. The complete sequence of the human KCNH2 gene described in GenBank (Accession Number NG 008916.1) was used as the reference.

### 4.3 SCN5A sequencing analysis

The results of the molecular evaluation of the *SCN5A* gene are presented in Table 5. All coding exons of this gene were analyzed. We were able to identify 7 polymorphisms of the *SCN5A* gene in our population (Table 2). The variants detected for this gene were: c.87A>G (p.A29A) in exon 2 of cases #2, #4, #5 and #6; c.1673A>G (p.H558R) in exon 12 of cases #3 and #4; c.5454T>C (p.D1818D) in exon 28 of case #2 and #4 and c.5841C>T (p.I1947I) in exon 28 of case #4; c.100C>T (p.R34C) in exon 2 of case #4 and c.537C>T (p.R179R) in exon 5 of case #6 and pE1061E in exon 17 in all cases except case #4 (Fig. 4). All of them located at the intracellular domain of the channel (Fig. 5).

**Fig. 4.**
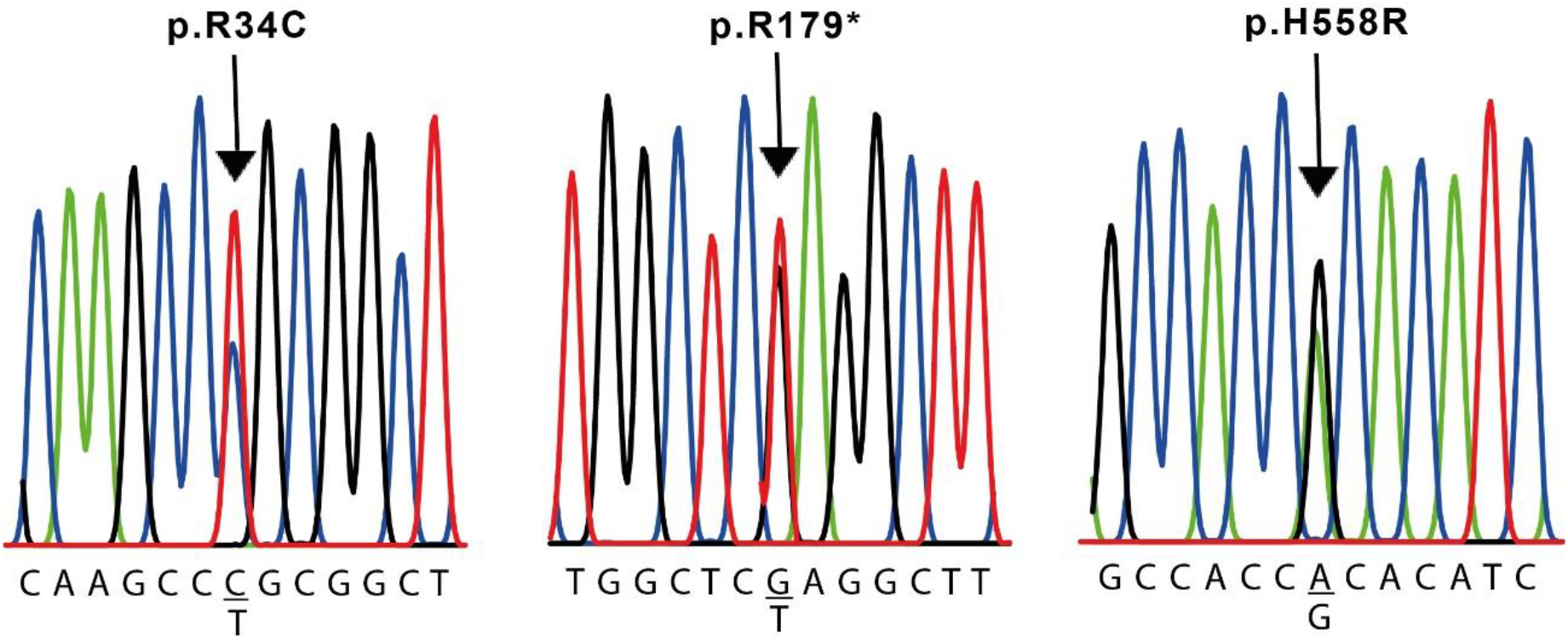
Nucleotide sequence of SCN5A variants in cases #4 and #6. The sequence is shown only for the sense strands of genomic DNA. The complete sequence of the human SCN5A gene described in GenBank (Accession Number NG 008934.1) was used as the reference.

**Fig. 5.**
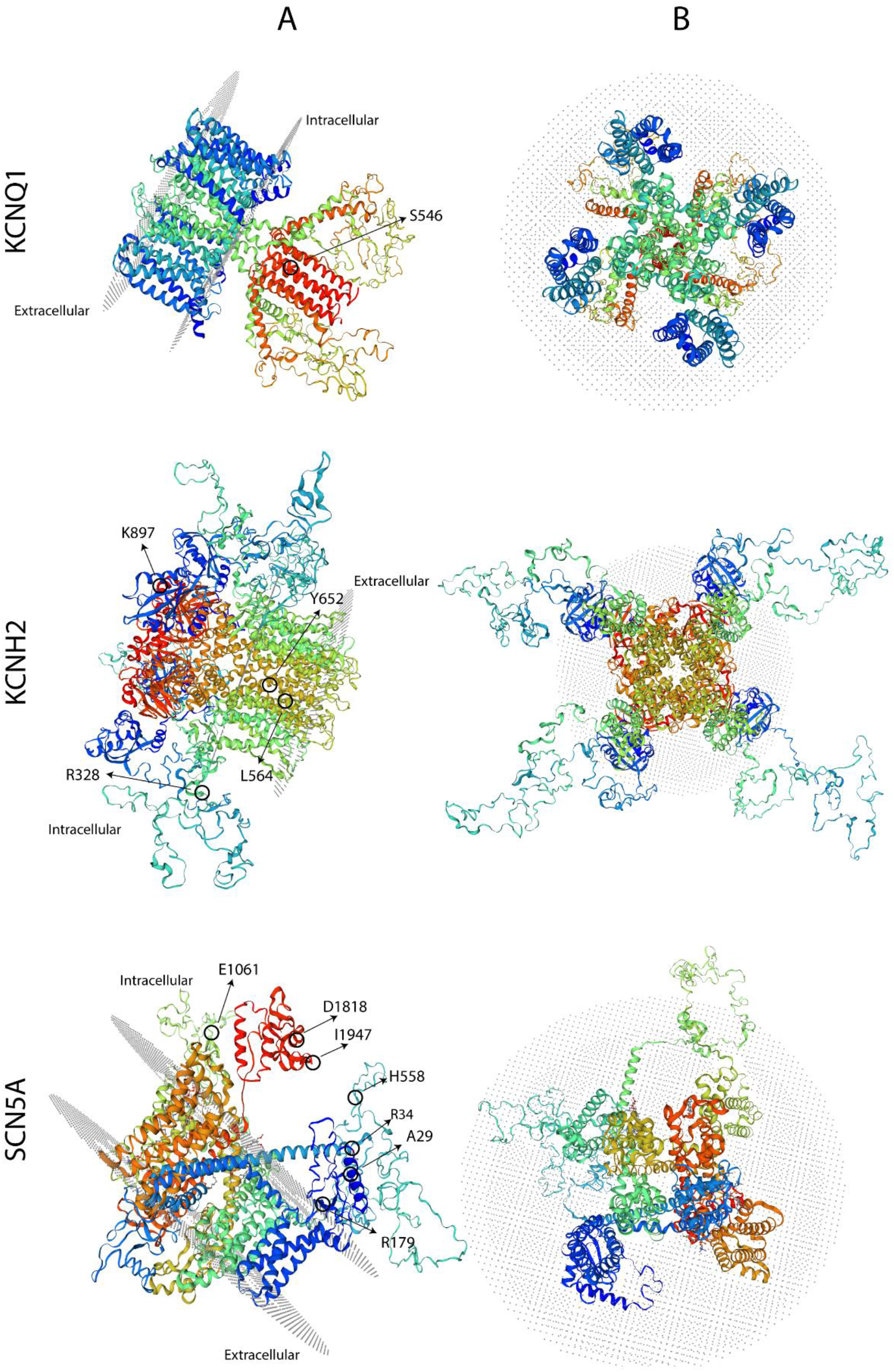
3D protein model with the mutations found in each channel. The Left panel shows the side view of the protein and the right panel, the upper (extracellular) view.

## 5. Discussion

### 5.1 *KCNQ1* gene

*KCNQ1* gene encodes the Kv7.1 potassium channel subunit responsible for slow rectifying K^+^ current (Iks) which requires four Kv7.1 subunits that co-assemble with the accessory KCNE1 subunits. KCNE1 subunits regulate channel activity but also modulate the affinity of Kv7.1 for phosphatidylinositol 4,5-bisphosphate (PIP2)^7^, which in turn controls channel gating^8^. Thus, the region of interaction between these two subunit types plays a significant role in the correct functioning of this channel in the human heart. Mutations in the *KCNQ1* gene can reduce or eliminate the Iks current in the human heart through different mechanisms, including prevention of subunit assembly, interruption of trafficking to the plasma membrane, or inhibiting their activation.

In this work, we found in exon 13 of the *KCNQ1* gene, the infrequent benign c.1638G>A variant in 2 out of the 6 patients and none of the controls. Its classification as benign is based on the fact that it is a silent mutation and therefore, implies no change in the amino acid sequence (p.S546S)^9^ (Table 5). However, although classified within this group, this variant has been found to be associated with various cardiac arrhythmias, such as LQT syndrome, atrial flutter, and atrial fibrillation, in European and Chinese individuals^9^. Moreover, in Chinese population has been associated with alterations in plasmatic lipid levels^10^. There is evidence indicating that synonymous mutations can affect the thermodynamic stability of mRNA secondary structures or affect splicing through phenomena such as exon skipping; therefore, synonymous mutations may not have neutral consequences in protein functional expression^11^. For example, if the stability of the mRNA is affected, the amount of protein synthesized could be reduced and, in the event of gene editing failures, a protein with an abnormal c-term domain could be generated. It should be noted as well that this variant is located in the domain that interacts with the accessory subunit KCNE1 (Fig. 5), which is necessary for proper channel function. This could then explain the functional abnormalities mentioned above^12^.

Noteworthy, in patient #5 we could not amplify exon 16 even with a second pair of primers which generate a bigger amplification product using two independent samples, suggesting a broad change in the DNA sequence for both alleles (Table 5). These changes could be a deletion in the portion of the sequence where the primers anneal, or an insertion big enough to not be amplified by our PCR conditions. This result may explain QT interval prolongation observed on the ECG scan. There is not much literature about exon 16 deletion but some authors have reported that young patients with deletions in exon 15 and 16 showed prolonged QTc^13,14^. Importantly, a small domain between residues 589 and 620 (i. e. within exon 16) in the KCNQ1 c-terminus may function as an assembly domain for KCNQ1 subunits^15^. This would imply that without this c-terminal domain, the KCNQ1 subunits would fail to assemble, thus preventing the formation of functional potassium channels. In this case then, partial or complete lack of exon 16 may therefore affect channel function through this mechanism.

### 5.2 *KCNH2* gene

The *KCNH2* gene encodes the ***α*** subunit of the HERG channel (Kv11.1). Functionally, this subunit co-assembles with the KCNE2 helper *β*-subunit to activate the rapid component of the delayed rectifying K^+^ current (Ikr) in the human heart during phases 2 and 3 of the cardiac action potential^16^. Mutations in *KCNH2* are responsible for the prolongation of the QT interval in a condition typified as LQT2, which leads to ventricular arrhythmia and, increases the risk of sudden death. The majority of mutations reported to date result in loss of function. Suggested mechanisms include interference with proper folding, assembly, or trafficking to the membrane^17^, defective activation/inactivation, loss in permeation selectivity and dominant-negative suppression of the function of wild-type channels^18^.

In the screening of the *KCNH2* gene of our population, we detected different types of variants in different exons. Two of them were silent without aminoacid changes. The silent variant c.1956T>C (p.Y652Y) in exon 8, was detected in 84% of our cases and none of the controls. This variant has been reported with a high frequency (0.67) in the European population^9^. Although this variant is silent, and this position seems to be important for the correct folding of the channel in the endoplasmic reticulum, changes in this position are associated with defects in channel trafficking and lead to LQT2 syndrome development^19^. The other silent variant found was c.1692A>G (p.L564L) which, is also classified as benign^20,21^.

The c.982C>T variant (p.R328C), which has been reported with a low frequency, falls into the pathogenic box^10^. Although this variant does not reduce the Ikr or alters channel function^22^, it has been associated to changes in conductance when is co-expressed with the *KCNQ1* mutation R591H^23^. Case #3 presents the R328C variant but not the R591H. However presents another variant in the *SCN5A* gene which could be responsible for the observed ECG changes (Table 5).

The second variant causing a change in the protein amino acid sequence is c.2690A>C (p.K897T). It is classified as benign but is also associated with channel kinetic changes, hERG expression and QT prolongation^24,25^. Also, some authors show an association between this variant and Atrial Fibrilation^26^. Besides, this variant acquires a pathogenic profile when it is associated to *KCNQ1* polymorphisms^12,24^. According to the literature, almost half of the suspected LQT2-causing mutations are missense mutations, and functional studies suggest that about 90% of these mutations disrupt the intracellular transport or trafficking of the *KCNH2*-encoded Kv11.1 channel protein to the cell surface membrane^27^. K897T is present in cases #2 and #6 togheter with silent variants in KCNQ1 and SCN5A genes (Table 5).

### 5.3 *SCN5A* gene

The *SCN5A* gene encodes the ***α*** subunit of the cardiac sodium channel (Nav1.5). LQT3 is generally associated with gain-of-function mutations in this channel. This leads to increased sodium entry into the heart muscle cells, causing it to take longer to recharge and thus prolonging the QT interval. The *SCN5A* gene can also undergo mutations that lead to loss of function of the sodium channel. These usually result in lower expression levels or translation into aberrant proteins that cause another type of electrical disease^28^.

Among the cases we analyzed, we detected several silent variants: p.A29A, p.E1061E, p.D1818D and p.I1947I and 3 disturbing ones: c.100C>T (p.R34C), c. 535C>T (p.R179*), and c.1673A>G (p.H558R). The silent variants were classified as benign since they produce no changes in the amino acid sequence^20,29–31^. R34C is a frequent polymorphism of this gene and is a relatively common in the black population, although it has also been detected among Hispanics^32^. c.537C>T (p.R179*) variant in *SCN5A* has been previously reported in individuals with Brugada syndrome and with long QT syndrome^33–35^. This variant leads to a premature termination codon at position 179, which is expected to result in an absent or non-functional protein. Although additional studies are required to fully establish its clinical significance, the R179* variant is classified as pathogenic. This variant is present in case #6, in heterozygouscity. This patient, also present the K897T in exon 11 of *KCNH2* gene, among other silent variants in the 3 studied genes. Problably the combination of all this DNA changes affects its cardiac function and prolong the QT interval. H558R is a common polymorphism present in 20-30% population and is associated with a prolonged QTc interval, which causes a change in the primary sequence. Some studies have reported it as a pathogenic mutation^32,36^, or associated with aberrations in the control of intracellular protein trafficking^24^, although others suggested that is a non-pathogenic polymorphism^20^ (Table 3). R558-containing sodium channels are wild-type–like except when co-expressed with the alternatively spliced transcript containing Q1077 whereby the channels are attenuated significantly^33^.

## 6. Conclusion

This is the first genetic testing of LQTS patients carried out in Argentina. Then, this study represents a kick-start for a systematic survey of the Argentinian population which is characterized by its heterogeneous genetic background.

Our analysis enabled us to detect several benign genetic variants and pathological changes in our population’s *KCNQ1, KCNH2*, and *SCN5A* genes. 5 cases present variations in the evaluated genes that may correlate with ECG long QT interval and clinical manifestations. The importance of detecting variants associated with alterations in the physiology and function of cardiac ion channels is highlighted, since they can create a vulnerable substrate that, in the presence of specific triggers such as IKr blockers, can precipitate life-threatening ventricular arrhythmias. Therefore, genetic analysis of suspected cases can provide valuable information to make an appropriate diagnosis and provide genetic counseling to relatives as well as to address the best treatment in both.

## Data Availability

All data produced in the present study are available upon reasonable request to the authors

## Funding

This work was supported by the VT38-UNS8614 grant from the Ministry of Education to G.S. and a grant from Universidad Nacional del Sur (PGI 24/B269) to E.A.

## Conflicts of interest

The authors declare that there is no conflict of interest.

## Acknowledgements

We would like to thank to the laboratory of the *Hospital Privado del Sur* and *IACA Laboratorios* for their support on DNA samples.

## Notes

### Competing Interest Statement

The authors have declared no competing interest.

### Funding Statement

This study was funded by Ministerio de Educacion VT38-UNS8614 and PGI 24/B269 and PGI 24/B296

### Author Declarations

Ethics committee/IRB of Hospital Municipal de Agudos Dr. Leonidas Lucero gave ethical approval for this work

